# Supporting the resilience and retention of frontline care workers in care homes for older people: A scoping review and thematic synthesis

**DOI:** 10.1101/2020.09.05.20188847

**Authors:** Lucy Johnston, Cari Malcolm, Lekaashree Rambabu, Jo Hockley, Susan D Shenkin

## Abstract

The COVID-19 pandemic has reinforced the need to ensure that strategic and operational approaches to retain high quality, resilient frontline care home workers, who are not registered nurses, are informed by context specific, high quality evidence. We therefore conducted this scoping review to address the question: What is the current evidence for best practice to support the resilience and retention of frontline care workers in care homes for older people?

MEDLINE, PubMed, PsycINFO, Embase, MedRxiv, CINAHL, ASSIA, Social Science Premium were searched for literature published between 2010 and 2020. The search strategy employed combinations of search terms to target frontline care workers in care homes for older people and the key concepts relevant to resilience and retention were applied and adapted for each database.

Thirty studies were included. Evidence for best practice in supporting the resilience and retention specifically of frontline care workers in care homes is extremely limited, of variable quality and lacks generalisability. At present, it is dominated by cross-sectional studies mostly from out with the UK. The small number of intervention studies are inconclusive.

The review found that multiple factors are suggested as being associated with best practice in supporting resilience and retention, but few have been tested robustly. The thematic synthesis of these identified the analytical themes of – Culture of Care; Content of Work; Connectedness with Colleagues; Characteristics and Competencies of Care Home Leaders and Caring during a Crisis.

The evidence base must move from its current state of implicitness. Only then can it inform intervention development, implementation strategies and meaningful indicators of success. High quality, adequately powered, co-designed intervention studies, that address the fundamentally human and interpersonal nature of the resilience and retention of frontline care workers in care homes are required.

## Background and context

The detrimental effects the COVID-19 pandemic had on the mental health of those who worked on the frontline through the height of the crisis has been quickly and well documented (Cullen, W. et al., 2020; Cabello, I.R., et al., 2020). Their experiences have focussed attention on the need to protect the psychological wellbeing of frontline health and social care workers globally.

To a large extent, however, much of the early and immediate support, resources, public attention and mitigation work was targeted at hospital based workers. As the severity of the impact of the pandemic on care homes became more evident, the critical need to support both residents and staff gained much needed impetus. This delayed focus was symptomatic of wider structural problems of an often overlooked and undervalued workforce (McGilton, K. et al., 2020; Devi, R. et al., 2020b).

The care home sector began 2020 already under considerable pressure, with little pre-preparedness for the additional demands of managing the pandemic on top of the challenges they were already facing. (Scottish Care, 2019; McGilton, K. et al, 2020). Ongoing recruitment and retention challenges (Oung, C. et al., 2020; Chen, HL, et al., 2012) will be exacerbated.

The vast majority of care home staff with a responsibility for providing direct care to residents are not registered nurses. We refer to this staff group as frontline care workers (FCWs). FCWs may be at greater risk of burnout given a number of factors, such as long and unsocial working hours, low pay and status, and the increasingly demanding physical and emotional nature of their work (VonDras et al. 2009; Health Foundation 2017; Dreher et al. 2018). Evidence suggests that the rate of turnover is greatest for FCWs (Donoghue et al. 2010; Rosen et al. 2011) who form the majority of staff within care homes. They have different training, skills and duties compared to the registered nurses they work alongside. Moreover, in contrast to registered nurses, FCWs are less likely to have connections to professional bodies or organisations. The impact of the recent COVID-19 pandemic has intensified the need to ensure this vital frontline care workforce is supported to build resilience, avoid burnout and remain in their roles delivering quality and compassionate care to older people.

However, the available evidence to inform best practice in supporting resilience and retention for frontline care workers in care homes is limited and of variable quality (Social Care Institute for Excellence, 2011). Evidence reviews of staff resilience conducted in response to the pandemic, focus on hospital based workers or all health and social care workforce (Heath, et al. 2020; Muller et al, 2020 and Pollock et al., forthcoming). The resulting broad nature of the developing evidence base cannot be transferred or generalised readily to care homes as they may not address sufficiently the ‘unique’ (University College London 2020), ‘special’ (Devi, et al., 2020a) and multi-faceted context of care homes (Social Care Institute for Excellence, 2011; Muller et al, 2020) nor those of the staff who work in them.

The COVID-19 pandemic has reinforced the need to ensure that strategic and operational approaches to retain high quality, resilient frontline care home workers, who are not registered nurses, are informed by specific, quality evidence. We therefore conducted this scoping review to address the question: What is the current evidence for best practice to support the resilience and retention of frontline care workers in care homes for older people?

## Methods

### Scope and definitions used

For purposes of this review, we use the term ‘resilience’ as a way of conveying not only the specific concept of resilience in itself (Scoloveno, 2016), but also of burnout, work-related stress, and psychological and mental health and wellbeing. Retention encompasses turnover rates, absenteeism, duration of employment and reported staff intentions to leave their job. There is evidence to support that building resilience amongst health care staff may be protective in avoiding burnout and thus in helping to retain staff in their roles (Cope et al. 2016; Badu et al. 2020; Delgado et al. 2020).

This review focusses only on those staff within care homes who have responsibility for providing direct care to residents, but are not registered nurses. We refer to them as frontline care workers (FCWs).

### Search Strategy

An initial search plan was developed (LJ, CM). A senior subject specialist librarian (SM) further developed and refined the search strategy and carried out the electronic database searches. Different combinations of search terms to capture FCWs in care homes for older people (Burton, J. et al., 2017) and the key concepts relevant to resilience and retention were applied and adapted for each database as necessary.

Eight databases (MEDLINE, PubMed, PsycINFO, Embase, MedRxiv, CINAHL, ASSIA, Social Science Premium) were searched for literature from 2010. Grey literature was located by applying the same search strategy principles. Internet searches of Google, Google Scholar and OpenGrey were undertaken. The websites of organisations and networks pertinent to health and social care were searched as were two COVID-19 specific sites – LitCOVID (NLM) and the WHO COVID-19 database. The search was undertaken in early June and repeated on July 16^th^ 2020 to ensure emerging evidence was captured.

The results of the database search were screened for relevance by reviewing the title and abstract. This was conducted independently by three members of the project team [SM, LJ, CM], and resulted in the initial inclusion of 222 papers. Full text versions of these papers were accessed and reviewed by three reviewers separately (LJ, CM and LR). Papers and publications were included if they met the criteria detailed in Table 1. Where there was no consensus for inclusion/exclusion a final decision was made by LJ (n = 3). This process resulted in the inclusion of 29 papers. The reference lists of the included articles were hand searched for further studies meeting the inclusion criteria and resulted in one additional paper. The PRISMA flowchart (figure 1) illustrates the search strategy and paper selection process.

**Table 1.** Inclusion and Exclusion Criteria.

| Inclusion | Exclusion |
| --- | --- |
| Published in English | Study protocols |
| Published between 2010-2020 | Reporting <u>only</u> on prevalence/measurement of resilience or retention |
| Setting is care homes for older people | Other residential settings, for example setting for physical/learning disabled adults |
| Provides evidence of practice based approaches to resilience and/or retention and explicitly states that it is of relevance to care home staff who provide direct care to residents | Evidence concerning resilience or retention which only includes or is only of relevance to registered nursing staff within care homes |
| Reports on findings or outcomes from evaluations of pilots, initiatives, activities, tests of change, QI programmes undertaken in care homes for older people | Discussions of conceptual frameworks or theoretical models of resilience and/or retention |

**Figure 1.**
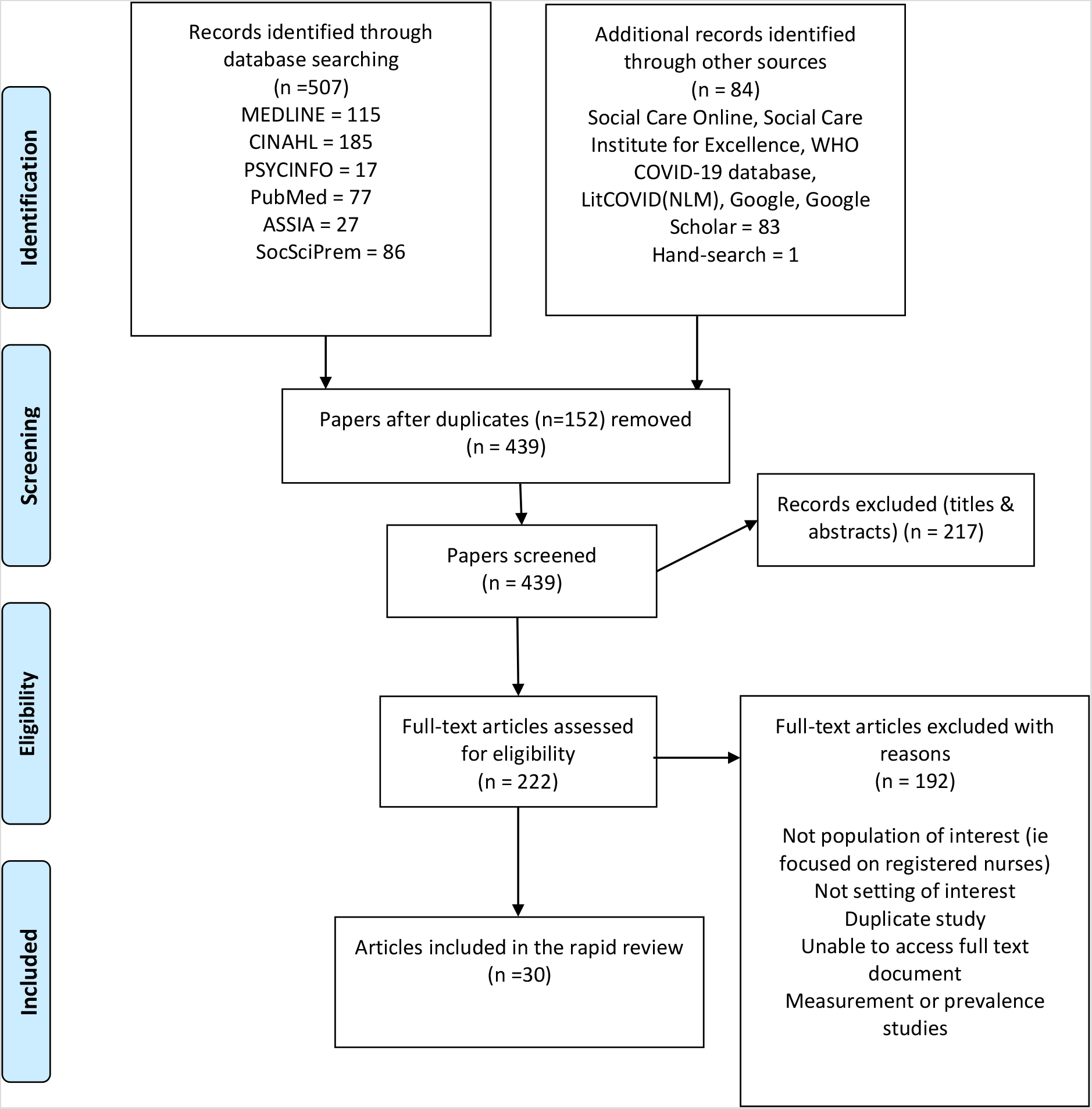
PRISMA flowchart illustrating the search strategy and paper selection process.

Given the targeted nature and emphasis of the review, we placed no restrictions on paper type. A decision was made not to exclude any source on the grounds of their ‘quality’. In accordance with scoping review methodology and given both the wide range of study designs included and the limited timeframe in which to undertake this review, a quality appraisal of the evidence was not undertaken (Arksey & O’Malley 2005; Levac et al. 2010; Tricco et al. 2018). Extracted data included: author(s), publication year, country of origin, study/paper design or methodology, aim, indicators and measures or resilience and/or retention, participants, findings and key recommendations.

### Thematic synthesis

Thematic synthesis of the results of the scoping review, adopting the three-stage method set out by Thomas and Harden (2008), was undertaken. Firstly, LR coded the extracted data, organising these into descriptive themes. A number of discussions between reviewers (LJ, LR and CM), were held. Discussion focussed on explicating the ‘meaning’ of the descriptive themes as they related to the review question and drawing out similarities, dissimilarities and patterns. Informed by this, LJ undertook the third and final stage of thematic synthesis and developed five analytical themes to ensure our findings went ‘beyond’ description and generated new insight and explanations (Thomas & Harden 2008). These were discussed and further refined by the whole team (LJ, CM, LR, JH and SS).

## Results

### Characteristics of included papers

General characteristics of the included papers are outlined in Table 2.

**Table 2.** Characteristics of included papers (n = 30).

| Source<br>(year of<br>publication) | Origin | Paper type/<br>Study design | Aim /Objectives | Setting | Participants | Primary<br>Focus |
| --- | --- | --- | --- | --- | --- | --- |
| Barbosa et al.<br>(2015a) | Portugal | Systematic<br>review | To assess the impact of PCC<br>approaches on stress, burnout,<br>and job satisfaction of staff caring<br>for people with dementia in<br>residential aged care facilities. | Residential<br>aged care<br>facilities<br>(dementia) | Direct Care Workers (DCWs) –<br>included nursing assistants/aides,<br>personal care attendants,<br>attendant care workers, personal<br>assistants, or frontline staff | Resilience |
| Barbosa et al.<br>(2015b) | Portugal | Pretest-Posttest<br>control group | To assess the effects of a PCC-based psychoeducational<br>intervention on direct care<br>workers' stress, burnout and job<br>satisfaction. | Residential<br>aged care<br>facilities<br>(dementia) | DCWs (n=58) - experimental<br>group (n=27) and control group<br>(n=31) | Resilience |
| Beck et al.<br>(2015) | Sweden | Pretest-Posttest<br>control group | To investigate the effects of an<br>intervention that applies a<br>palliative care approach in<br>residential care upon nurse<br>assistants' level of strain, job<br>satisfaction, and view of<br>leadership. | Care homes | Nurse Assistants (n=185) -<br>experimental group (n=75) and<br>control group (n=110) | Resilience<br>and<br>retention |
| Berridge et al.<br>(2018) | USA | Cross-sectional<br>survey | To examine whether staff<br>empowerment practices common<br>to nursing home culture change<br>are associated with certified<br>nursing assistant (CNA) retention. | Nursing homes | Nursing Home Administrators<br>(n=2,034) with reference to their<br>Certified nursing assistant staff | Retention |
| Berridge et al. (2020) | USA | Cross-sectional survey | To examine the relationship between nursing assistant (NA) retention and a measure capturing nursing home leadership and staff empowerment. | Nursing homes | Nursing Home Administrators (n=1,386) with reference to their Certified nursing assistant staff | Retention |
| Berta et al. (2018) | Canada | Cross-sectional survey | To examine the relationships among perceptions of the work environment, work attitudes, and work outcomes of HSWs engaged in providing care to older Canadians in long-term care and community care settings in Ontario, Canada. | Nursing homes | Health support workers (HSWs) (n=460) | Retention |
| Bethell et al. (2018) | Canada | Cross-sectional survey | To examine the association between supervisory support and intent to turn over among personal support workers (PSWs) employed in long-term care homes in Ontario, Canada. | Care homes | Personal support workers (PSWs) (n=5,513) | Retention |
| Boerner et al. (2017) | USA | Qualitative interviews | To investigate staff, institutional, patient, and grief factors as predictors of burnout dimensions among direct care workers who had experienced recent patient death; determine which specific aspects of these factors are of particular importance; and establish grief as an independent predictor of burnout dimensions. | Nursing home | Certified nursing assistants (CNAs) (n=143) | Resilience |
| Braedley et al. (2018) | Canada | Qualitative interviews (secondary analysis of existing dataset) | To explore relationships between long term residential care nursing staff's psychological health and wellbeing and working conditions that include work overload, low worker control, disrespect and discrimination. | Care homes | Registered Nurses (n=20); Licensed Practical Nurses (n=20); Continuing Care Aides (n=44); Social workers (n=3) | Resilience |
| Caspar et al. (2020) | Canada | Institutional Ethnography (IE) | To explore how the social organisation of work influences the quality of work-life and care delivery in long term care homes. | Care homes | Resident care aides (RCAs) (n=42) | Resilience |
| Castle (2013) | USA | Cross-sectional survey | To examine the associations with consistent assignment of nursing aides with low turnover and low absenteeism. | Nursing homes | Nursing Home Administrators for n=3,941 facilities with reference to their Nurse aide staff members | Retention |
| Choi & Johantgen (2012) | USA | Cross-sectional survey | To examine the relationships of work-related and personal factors to CNA job satisfaction and intent to leave. | Nursing homes | Certified nursing assistants (CNAs) (n=2,254) | Retention |
| Dreher et al. (2019) | USA | Pretest-Posttest single group design, mixed methods approach | To increase CNA retention through an evidence-based education training program on compassion fatigue awareness and multiple self-care skill strategies. | Nursing homes | Certified nursing assistants (CNAs) (n=45) | Resilience and retention |
| Ericson-Lidman & Ahlin (2017) | Sweden | Pretest-Posttest single group | To compare assessments of stress of conscience, perceptions of conscience, burnout, and social | Care homes | Registered Nurses (n=5)<br>Nurse assistants (n=24) | Resilience |
|  |  | design, using surveys | support among health care personnel (HCP) working in municipal residential care of older adults, before and after participation in a participatory action research (PAR) intervention aiming to learn to constructively deal with troubled conscience. |  |  |  |
| Fukuda et al. (2018) | Japan | Cluster, quasi-randomised, controlled comparative trial | To evaluate the effectiveness of educational intervention using printed educational material for reducing distress induced by behavioural and psychological symptoms of dementia among caregivers working at facilities without medical specialists and/or registered nurses. | Residential aged care facilities (dementia) | Intervention Group (n=185 staff)<br>Control Group (n=172 staff)<br>Staff groups included care workers, nurses, occupational therapists, clinical psychologists.<br>Breakdown of staff numbers not provided in paper but care workers made up 60% of the total sample. | Resilience |
| Gaudenz et al. (2019) | Switzerland | Cross-sectional survey (secondary analysis of existing dataset) | To evaluate the prevalence and variability of nursing home care workers' intent to leave. | Nursing homes | Care workers (n=3,949) including Registered Nurses, Licensed Practical Nurses, Certified Nursing Assistants, Nurse Aides | Retention |
| King et al. (2013) | Australia | Cross-sectional survey | To examine the effects of worker satisfaction, worker characteristics, work conditions, and workplace environment on intention to leave. | Care homes | Personal care assistants (n=4,316) | Retention |
| Lane & McGrady (2018) | USA | Narrative review and exploratory study | To determine how the CMS Emergency Preparedness Checklist contributes to organisational resilience by a) identifying the adaptive capacity and planning factors addressed by the CMS Emergency Preparedness Checklist, and b) identifying the adaptive capacity and planning factors not addressed by the CMS Emergency Preparedness Checklist.<br>To recommend tools and processes to improve adaptive capacity and planning for long-term care facilities. | Nursing homes |  | Resilience |
| McNeil et al. (2019) | Australia | Cross-sectional survey | To examine the impact of personal resilience on the wellbeing of care workers and how perceptions of the quality of care provided and the social climate in the organisation influences this relationship. | Care homes | Care workers (n=140) | Resilience |
| Moss & Meyer (2014) | UK | Research briefing | To summarise the key findings of a research review on 'keeping the workforce fit for purpose' undertaken. | Care homes |  | Resilience |
| Nakanishi & Imai (2012) | Japan | Cross-sectional survey | To examine job role quality relating to intention to leave current facility and to leave profession among direct care workers in residential facilities for elderly in Japan. | Nursing homes | Direct care workers (n=3,527) | Retention |
| Rajan & Mckee (2020) | UK | Online pilot survey | To report how the lived experiences of care home providers can provide important insights that inform a whole system response that will be required to prevent future avoidable fatalities in care homes in the event of a second wave of infections. | Care homes | Care Home Managers (n=42) and Directors (n = 35) | Resilience |
| Schwendimann et al. (2016) | Switzerland | Cross-sectional survey | To describe job satisfaction among care workers in Swiss nursing homes and to examine its associations with work environment factors, work stressors, and health issues. | Nursing homes | Care workers (n=4,145) including Registered Nurses, Licensed Practical Nurses, Certified Nursing Assistants, Nurse Aides | Retention |
| Scottish Care (2019) | UK | Report | To understand more about the mental health of people living in care homes and accessing care at home and housing support services and of the social care workforce. | Care homes, care at home and housing support service | All front line social care staff including, but not limited to, support workers | Resilience |
| Wallin et al. (2012) | Sweden | Cross-sectional survey | To investigate job satisfaction and explore associated variables | Residential care facilities | Nursing assistants (n=225) | Resilience |
|  |  |  | among nurse assistants working in residential care. | for older people (general or dementia) |  |  |
| Yeatts et al. (2018) | USA | Cross-sectional survey | To identify factors associated with burnout among DCWs in nursing homes. | Nursing homes | Direct care workers (DCWs) (n=410) | Resilience |
| Yeatts et al. (2010) | USA | Cross-sectional survey | To examine the relationship between the certified nurse aides' (CNAs) perception that 'training is always available when needed' and the CNAs performance, turnover, attitudes, burnout and empowerment. | Nursing homes | Certified nurse aides (n=359) | Resilience and retention |
| British Geriatrics Society (2020) | UK | Good Practice Guide |  | Care homes |  | COVID-19 Guidance |
| University College London (2020) | UK | Guidance document |  | Care homes |  | COVID-19 Guidance |
| World Health Organisation (WHO) (2020) | International | Guidance document |  | Long-term care services |  | COVID-19 Guidance |

### Country of origin

Nine papers originated from the USA (Berridge et al. 2018; Berridge et al. 2020; Boerner et al. 2017; Castle 2013; Choi & Johantgen 2012; Dreher et al. 2019; Lane & McGrady 2018; Yeatts et al. 2010; Yeatts et al. 2018), five from the UK (Moss & Meyer 2019; Scottish Care 2019; British Geriatrics Society 2020; Rajan & Mckee 2020; University College London 2020), four from Canada (Berta et al. 2018; Bethell et al. 2018; Braedley et al. 2018; Caspar et al. 2020), three from Sweden (Beck et al. 2015; Ericson-Lidman & Ahlin 2017; Wallin et al. 2012), and two from Switzerland (Gaudenz et al. 2019; Schwendimann et al. 2016), Japan (Fukuda et al. 2018; Nakanishi & Imai 2012), Portugal (Barbosa et al. 2015a; Barbosa et al. 2015b), and Australia (King et al. 2013; McNeil et al. 2019). One paper was published by an international organisation (World Health Organisation 2020).

### Setting and participants

Care homes for older people were referred to in the papers by a range of terms such as nursing homes, residential aged care facilities and long term care facilities. Job titles included, direct care workers (DCWs), certified nursing assistants (CNAs), and licensed practical nurses (LPNs).

Almost half the papers (14) had only FCWs as the participants (Barbosa et al., 2015b; Beck et al. 2015; Berta et al. 2018; Bethell et al. 2018; Boerner et al. 2017; Caspar et al. 2020; Choi & Johantgen 2012; Dreher et al. 2019; King et al. 2013; McNeil et al. 2019; Nakanishi & Imai 2012; Wallin et al. 2012; Yeatts et al. 2010; Yeatts et al. 2018), and six papers included all care home staff, comprising both registered nurses <u>and</u> what we have termed as FCWs (Braedley et al. 2018; Ericson-Lidman & Ahlin 2017; Fukuda et al. 2018; Gaudenz et al. 2019; Schwendimann et al. 2016; Scottish Care 2019). Three papers included care home administrators/managers (Berridge et al. 2018, 2020; Castle 2013) and in one paper, participants were the managers and directors of care homes (Rajan & Mckee 2020).

### Paper Type/Study Design

The majority (n = 25) were empirical research, mostly cross-sectional survey studies (Berridge et al. 2018, 2020; Berta et al. 2018; Bethell et al. 2018; Castle 2013; Choi & Johantgen 2012; Gaudenz et al. 2019; King et al. 2013; McNeil et al. 2019; Nakanishi & Imai 2012; Rajan & Mckee 2020; Schwendimann et al. 2016; Wallin et al. 2012; Yeatts et al. 2010, 2018). Only five papers reported pre-test/post-test evidence (Barbosa et al. 2015b; Beck et al. 2015; Dreher et al. 2019; Ericson-Lidman & Ahlin 2017; Fukuda et al. 2018). Of these, one was a quasi-randomised comparative trial (Fukuda et al. 2018).

Five papers present evidence derived from qualitative approaches - interviews with individuals or group discussions (Boerner et al. 2017; Braedley et al. 2018; Scottish Care 2019), institutional ethnography (Caspar et al. 2020).

One systematic review (Barbosa et al. 2015a), a narrative review (Lane & McGrady, 2018) and a research briefing containing a review of relevant literature (Moss & Meyer 2014) were also included. Three papers were COVID-19 specific guidance or good practice documents (British Geriatrics Society 2020; University College London 2020; World Health Organisation 2020).

Ten papers focussed primarily on the retention of care home staff (Berridge et al. 2018, 2020; Berta et al. 2018; Bethell et al. 2018; Castle 2013; Choi & Johantgen 2012; Gaudenz et al. 2019; King et al. 2013; Nakanishi & Imai 2012; Schwendimann et al. 2016) and 13 on staff resilience (Barbosa et al. 2015a, 2015b; Beck et al. 2015; Boerner et al. 2017; Braedley et al. 2018; Caspar et al. 2020; Ericson-Lidman & Ahln 2017; Fukuda et al. 2018; McNeil et al. 2019; Moss & Meyer 2014; Scottish Care, 2019; Wallin et al. 2012; Yeatts et al. 2018).

Organisational resilience was the focus of a 2018 report from the USA reviewing aspects of nursing home resilience in relation to emergency preparedness (Lane & McGrady, 2018) but additionally discussed individual staff resilience ‘competencies’ and indicators. The work of Rajan & Mckee (2020) was published during the COVID-19 pandemic, reporting on the key workforce challenges care homes were facing including the impact on staff morale, mental health and wellbeing.

Whilst the three COVID-19 guidance papers covered managing the pandemic within care homes, they included recommendations and/or principles for supporting care home staff mental health and well-being (British Geriatrics Society 2020; University College London 2020; World Health Organisation 2020).

Three papers (Dreher et al., 2019; Beck et al., 2015; Yeats et al., 2010) explicitly addressed both topics of interest to this review by linking aspects of resilience to retention. Dreher et al (2019) investigated whether retention would improve if awareness of compassion fatigue and self-care strategies amongst staff was increased. Yeatts et al (2010) utilised data from a larger study to explore staff perceptions on how the training available to FCWs affected their performance, turnover, attitudes, burnout, and empowerment. Beck et al (2015) investigated the effects on nurse assistants’ work situation of applying a palliative care approach.

### Intervention studies

Five papers reported evaluations of an educational intervention on a range of indicators of resilience and retention (Table 3). Increased retention rates were observed at one and four months following attendance at a 90-minute educational programme addressing self-care skills and awareness of compassion fatigue (Dreher et al. 2019).

**Table 3:**
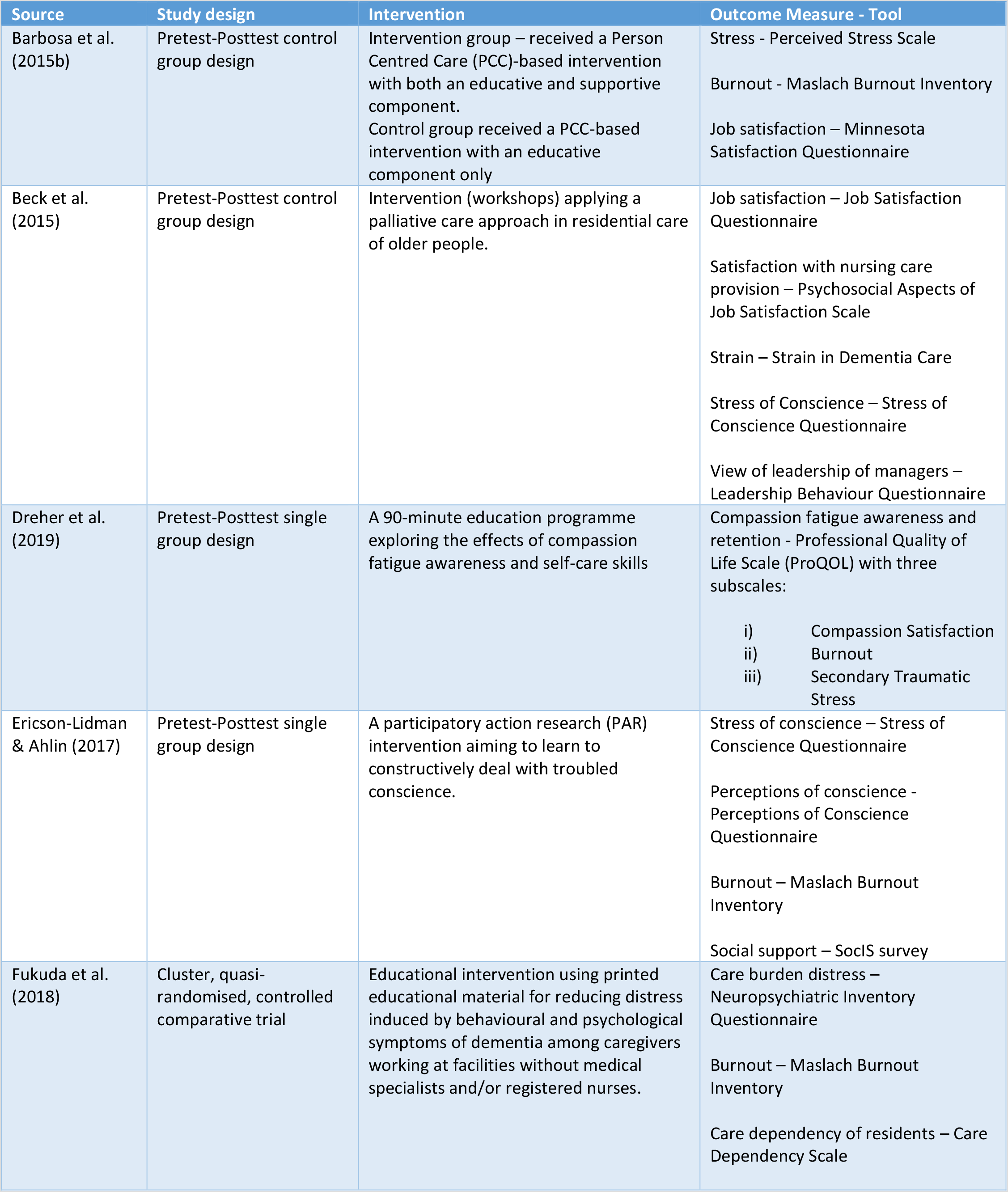
Intervention Studies included with outcome measure used.

Dreher et al. (2019) also measured ‘burnout’ using the Maslach Burnout Inventory (MBI). They found that the non-parametric test (Kruskal-Wallis) showed a significant reduction in burnout score post-intervention, however conducting a parametric test ANOVA did not find a statistical significance in burnout scores in the same study. No statistically significant reduction score post-intervention was found by two other intervention studies (Fukuda et al. (2018), Ericson-Lidman & Ahlin (2017).

When a ‘supportive component’ was added to an educational intervention, researchers found a statistically significant reduction in the emotional exhaustion component of MBI, compared to those who received a purely educational intervention (Barbosa et al. 2015b). Qualitative analysis of perception and impact of the psychoeducational intervention on their work life showed that the experimental group experienced enhanced group cohesion, emotional management, and self-awareness (Barbosa et al. 2015b).

Overall, the results of the systematic review and the five intervention studies were limited and inconclusive. Of note was the marked heterogeneity in the outcome measures of resilience: 13 different inventories or measurement tools were used across the five studies.

### Predictors and indicators of resilience and/or retention reported in the evidence

Multiple predictors and indicators of resilience and retention were found within the papers. The factors investigated and reported within each paper covered those that were (a) hypothesised and then investigated; (b) found to influence or impact on; or (c) raised in the discussion section as possibly or potentially associated with resilience or retention of FCWs. These ranged from self-care behaviours of individual staff members (Dreher et al. 2017, University College London 2020) to, for example, the overall organisational environment and context within which these individual work (Yeatts et al. 2018; King Et al 2013; Moss & Meyer 2014; Lane & McGrady 2018 and University College London 2020). To a large extent, this multiplicity reflects the specific hypothesis/aims of the studies. The strength of association of these factors for improving resilience and retention cannot be determined sufficiently.

For example, the studies that examined leadership as a factor in resilience or retention included measures of stress of conscience, wellbeing, job satisfaction, and rates of staff turnover. Two studies with different participant groups – one with nursing home care workers and the other with nursing administrators – both reported a strong relationship between leadership and retention of staff (Gaudenz et al. 2019 and Berridge et al. 2020). Care workers with higher overall intention to leave reported lower leadership ratings (Gaudenz et al. 2019). Berridge and colleagues (2020), in their survey of nursing home administrators, reported greater leadership and staff empowerment levels were associated with high retention of nursing assistants.

In one large cross-sectional study, job satisfaction was found to increase four times with each point increase in leadership rating on a 4-point Likert-type scale (Schwendimann et al. 2016). Positive leadership was also reported to contribute to a low stress of conscience i.e. nursing assistants were better able to provide care that corresponded to their own conscience when there was better leadership (Wallin et al. 2015). One study found that leadership styles had a negative effect on nursing assistants’ wellbeing post-intervention and how this leadership was perceived by nursing assistants varied significantly over time (Beck et al. 2015).

### Analytical themes from the results of the review

Our thematic analysis provided further insight in relation to practice-based ways of supporting the resilience and of FCWs in care homes for older people. Five analytical themes were identified - Culture of Care; Content of Work; Connectedness with Colleagues; Characteristics and Competencies of Care Home Leaders and Caring during a Crisis. Table 4 shows how each included paper contributed to the development of each theme.

**Table 4:** Contribution of each paper to analytical themes

| Source | Content of Work | Connectedness with colleagues | Competencies/ Characteristics of Leaders | Culture of Care | Caring during a Crisis |
| --- | --- | --- | --- | --- | --- |
| Barbosa et al. | x |  |  |  |  |
| Barbosa et al. |  | x |  |  |  |
| Beck et al. |  |  | x |  |  |
| Berridge et al. | x | x |  | x |  |
| Berridge et al. |  |  | x | x |  |
| Berta et al. |  | x |  | x |  |
| Bethell et al. |  | x |  |  |  |
| Boerner et al. | x | x |  | x |  |
| Braedley et al. | x | x |  | x |  |
| Caspar et al. |  | x | x |  |  |
| Castle | x |  |  |  |  |
| Choi & Johantgen |  | x |  | x |  |
| Dreher et al. | x |  |  | x |  |
| Ericson-Lidman & Ahlin |  |  |  | x |  |
| Fukuda et al. | x |  |  | x |  |
| Gaudenz et al. |  | x | x | x |  |
| King et al. (2013) | x |  |  | x |  |
| Lane & McGrady |  | x | x | x | x |
| McNeil et al. | x |  |  | x |  |
| Moss & Meyer |  |  |  | x |  |
| Nakanishi & Imai | x | x |  | x |  |
| Rajan & McKee |  | x | x | x | x |
| Schwendimann et al. |  | x | x |  |  |
| Scottish Care |  | x | x | x |  |
| Wallin et al. | x |  | x | x |  |
| Yeatts et al. | x |  | x | x |  |
| Yeatts et al. |  |  |  | x |  |
| British Geriatrics Society |  | x |  |  | x |
| University College London |  | x | x | x | x |
| World Health Organisation |  | x |  |  | x |

### Culture of Care

This theme encompasses the culture of caregiving and the broader environment or climate within which FCWs work. It highlights the importance of FCWs being able to work in a climate that respects their role and the contribution they make to individual residents and to wider society. Seven papers discuss the way in which being respected, feeling respected and valued and receiving recognition can have a positive impact on staff resilience and retention (Yeatts et al. 2018; Choi & Johantgen et al. 2012; Gaudenz et al. 2019; King et al. 2013; Nakanishi & Imai 2012; Rajan & Mckee 2020 and Moss & Meyer 2014).

A number of papers in this review draw a specific connection between a care home’s culture of person-centred care (PCC) to resilience and retention. Wallin et al. (2012) discuss the positive benefits to staff of being able to provide good PCC and others investigate the relationship between PCC training and delivery to stress, burnout and job satisfaction (Barabosa et al. 2015 a; Barabosa et al 2015b).

Moss & Meyer (2014) and Boerner et al. (2018) suggest that, the way in which PCC is different from a task-orientated culture, offers ‘a psychological defence mechanism against anxiety’, giving have a ‘protective effect’ on FCWs. Berridge et al. (2018) suggests that the hours a FCW spends each day with residents is also important and the work of Castle (2018) that of staff being able to consistently work with the same residents. These two associated factors are also contributing to the Content of Work theme as is the role ‘job satisfaction’ can play in resilience and retention.

Seven papers propose that job satisfaction is a key factor in resilience and retention. Of these, five papers view it as an important factor in retaining staff (Berta et al. 2018; Choi & Johantgen 2012; Bethell et al 2018; King et al 2013; Beck et al 2015) and one in fostering staff resilience (Schwendimann et al. 2016). The seventh paper investigated determinants of job satisfaction (Wallin et al. 2012) and identified various aspects of work content and work climate/culture as being of importance, in addition to what they term ‘organisational and environmental support’.

### Content of Work

Content of Work theme encompasses the factors reported in the evidence that are related to *what* FCWs do in practice – that is their actual tasks, activities and jobs they undertake and as discussed above how satisfied staff are with this.

The overall design of work content is highlighted as being associated with resilience and retention (Yeatts et al 2018). How tasks are allocated will determine how much time staff spend with which residents, reported as a positive factor in staff retention by two papers (Castle 2018 and Berridge et al. 2018).

Nakanishi & Imai (2012) found that intention to leave was associated with the extent to which FCWs had discretion in how they used their skills and Braedley et al. (2018) identified that having autonomy of tasks was of importance. The degree of staff empowerment as an associated factor was reported in three papers (Berridge et al. 2018; Berridge et al. 2020; Lane & McGrady 2018) as was FCWs being involved in care decisions (Braedley et al. 2018). A lack of variety of work content/tasks and work content resulting in skills being underused were found to affect retention negatively (Nakanishi & Imai 2012 and King et al 2013).

### Connectedness with Colleagues

The extent and quality of a FCW’s connection and relationships with colleagues are associated with resilience and retention. At a high level, connectedness with colleagues is more commonly referred to as peer support and team working.

Having good, positive one-to-one relationships with work colleagues (Schwendimann et al. 2016; Nakanishi & Imai 2012; Gaudenz et al 2019; King et al 2013; Casper et al 2020) is viewed as being associated with resilience and retention. More specifically, three papers identify the absence of ‘conflict’ in these relationships as important (Gaudenz et al 2019; King et al 2013 and Schwendimann et al 2016) and four others the importance of a FCW’s relationships with their immediate supervisor (Bethell et al 2018; Choi & Johantgen 2012; Berta et al 2018 and Nakanishi & Imai 2012).

Team working was reported by eleven of the papers in the review (Barabosa 2015a; Berridge et al. 2018; Berridge et al. 2020; Braedly et al 2018; Casper et al 2020; Schwendimann et al. 2016; Rajan & Mckee 2020; University College London 2020; Gaudenz et al 2019; Scottish Care 2019 and WHO 2020). Reciprocity (Casper et al 2020) and communication (Braedley et al 2018) were specified as contributing to ‘good’ team working. No other detailed information was reported to better define what particular aspects of team working are most associated with resilience and retention. Three papers spoke of team working beyond staff groups as being of importance, indicating the value of wider multi-disciplinary or multi-sector teams (Scottish Care, 2019; University College London, 2020 and WHO, 2020).

### Characteristics and competences of leaders in care homes

Five studies reported on the relationship between leadership and resilience and retention. Three studies, all with different participant groups, reported a strong relationship between leadership and retention of staff (Gaudenz et al. 2019: Berridge et al. 2020; Schwendimann et al. 2016). Positive leadership was also reported to contribute to a low stress of conscience i.e. nursing assistants were better able to provide care that corresponded to their own conscience when there was better leadership (Wallin et al. 2015).

Within the included papers there is also some indication that in addition to skill/competencies, management/leadership ‘style’ is also important (Berridge et al. 2018 and Beck et al. 2015). Other papers highlight desirable characteristics of leaders. These include for example being compassionate (University College London 2020); positive (Wallin et al 2012); supportive (Choi & Johantgen 2012; Schwendimann et al. 2016; Bethell et al 2018; Boerner et al 2017; Berta et al 2018); visible (Rajan & Mckee 2020), inclusive and responsive (Casper et al 2020).

Bethell et al. 2018, Gaudenz et al. 2019 and Schwendimann et al. 2016 all suggest the need for leadership training for home managers and those in ‘middle management positions’. Two papers highlighted the need to ensure leaders possess the skills to embed good practice post training. (Yeatts et al 2010 and Beck et al 2015).

### Caring during a Crisis

Five papers were concerned with how care homes operate during a pandemic (Lane & McGrady 2018; WHO 2020; University College London 2020; British Geriatric Society 2020; and Rajan & Mckee 2020). The four themes previously outlined are also evident within the two reports and three guidance documents published during the COVID-19 pandemic. However, the importance of positive and supportive peer relationships and team cohesion are elevated (University College London 2020; British Geriatric Society 2020; Rajan & Mckee 2020; WHO 2020). Moreover, the critical role of care home leaders in supporting and facilitating care during a crisis is a priority (University College London 2020; Rajan & McKee 2020; Lane & McGrady 2018)

The need to ensure FCWs are aware of where to access support and provision of dedicated grief and bereavement support are also highlighted as of importance to staff support during a crisis (University College London 2020, WHO 2020)

## Discussion

This review found that the evidence for best practice in supporting the resilience and retention specifically of FCWs in care homes is extremely limited. The small number of intervention studies are inconclusive. Multiple factors reported as being associated with how best to support FCWs were identified. From this diffuse and dilute evidence base, our thematic synthesis distils important areas that warrant further exploration and research. The results of this review suggest that existing evidence provides insight into ‘promising’ avenues, but at present offers less in developing our understanding of how best to construct services and systems that can be implemented within care homes. The evidence base must move from its current state of implicitness to one of detailed explication. Only then can it inform intervention development, implementation strategies and meaningful indicators of success.

High quality, adequately powered and co-designed intervention studies are now required to determine which factors are of most importance, *how* they ‘work’ or ‘don’t work’ alone or in combination, and can be enhanced for positive effect. We need to for example, understand better (1) how the overall culture of care homes and an individual’s work content relates to job satisfaction and intention to stay. (2) the way in which FCWs interact, communicate and work together is both positively and negatively associated with retention and resilience and (3) the skills and approaches care home leaders have or need and the role of education and training.

Emerging as perhaps worthy of more intensive investigation are the potential of PCC as a protective mechanism for both resilience and retention and the more nebulous concepts of support and job satisfaction.

## Strengths and Limitations

This review purposefully examined only the evidence available for practice based resilience and retention support FCWs for older people. As such it does not cover other staff that are critical to the delivery of high quality care and the resilience and retention of FCWs – most importantly registered nurses. It has also excluded broader aspects of recruitment and retention such as pay, or demographic issues such as age and gender or geographical demographics that will affect the labour market. Only studies published in English were included as time and budget constraints did not allow for translation of papers.

Resilience is a wide and multi-faceted field and we make no claim to have utilised it in any great detail within this paper. However, it is a term in common use and used in this review to convey and include burnout, mental wellbeing, mental health, psychological wellbeing as it relates to being employed.

This rapid review is the first to our knowledge that focusses solely on FCWs in care homes. It addresses not only an under-researched staff group and provides much needed targeted review of available evidence as to how best they can be supported. It incorporates what was known pre-COVID and also what has been found to be of use during the pandemic for supporting resilience of FCWs in care homes. Although small and rapid the involvement of a specialist librarian and three independent reviewers are further key strengths of this work.

## Conclusion

This review sets out the evidence currently available for best practice in supporting a resilient workforce and retaining frontline care workers in care homes. The thematic synthesis has identified important areas that warrant further exploration and research within a very heterogeneous care service and workforce sector. Therefore the development of evidence based, best practice cannot just focus on what can be done differently in terms of new interventions, training or systems; but critically must address *how* and *where* (in what context) it is done. The fundamentally human and interpersonal nature of the resilience and retention of FCWs in care homes is highlighted by this review. This insight and perspective should inform future strategic and operational approaches to retain high quality, resilient frontline care home workers.

## Data Availability

Available by contacting the author(s)

## Acknowledgements

Sheena Moffat Edinburgh Napier University

